# Poultry Ownership in Urban Kenya is Associated with Increased Fecal Contamination in Household Soil

**DOI:** 10.1101/2023.09.16.23295449

**Authors:** Elana M. G. Chan, Jenna M. Swarthout, Benard Chieng, Sylvie Araka, John Mboya, Christine Imali, Angela R. Harris, Sammy M. Njenga, Amy J. Pickering

## Abstract

Animal feces can contain zoonotic enteropathogens capable of causing human diarrheal disease. Limited knowledge exists on domestic animal management in low-income urban settlements. We leveraged survey data and environmental samples collected from 120 urban Kenyan households to understand poultry husbandry practices and assess if household poultry ownership was associated with *Escherichia coli* contamination in stored water and soil. Fifty-five percent (n = 66) of households were in poultry-owning compounds, and 59.1% (n = 39) of these households reported poultry entering the household quarters. Among these 39 households, 53.9% (n = 21) kept poultry in the sleeping quarters of under-5 children. Household poultry ownership (49.2%, n = 59) was associated with increased *E. coli* concentrations in soil but not with *E. coli* prevalence in stored water. Poultry husbandry in urban settings may promote zoonotic disease transmission, and household soil may be an important transmission pathway for poultry-associated fecal contamination.

---

Diarrheal disease was the third leading cause of under-5 child mortality worldwide in 2017.^1^ Exposure to enteric pathogens in animal feces has been associated with diarrheal disease in humans, and roughly 60% of known infectious diseases can be transmitted zoonotically.^2,3^ Previous studies in low-income, rural households in Kenya, Bangladesh, and India detected *Escherichia coli*, a fecal indicator bacteria (FIB) of which some strains are diarrheagenic, originating from humans and domestic animals in household environments.^4–7^ In Dhaka, Bangladesh, FIB and a ruminant-associated bacterial target were identified in hand rinses and floor samples from households both owning and not owning ruminants, suggesting that fecal contamination from animals is widespread in low-income, urban settings.^8^ Urban settlements are characterized by overcrowding and less land area per household; animal rearing practices differ from rural areas and humans may have high contact with animals because there is less space to separate animals from living quarters or for animals to graze.^9^ As urban migration increases, there is a need to better understand animal husbandry practices and transmission pathways of zoonotic enteropathogens in densely populated areas. We used household survey data and environmental (soil, water) samples in urban Kenya to assess (i) poultry husbandry practices and (ii) whether household poultry ownership was associated with *E. coli* contamination in environmental samples.

Between June–August 2019, 120 eligible households including at least one under-5 child were enrolled from two subcounties in Nairobi, Kenya: Kibera (n = 64) and Dagoretti South (n = 56) (Figure S1). Systematic sampling efforts were made to ensure approximately half of the households were from poultry-owning compounds. For every compound enrolled with poultry on a street, another compound without poultry was enrolled. Only one household was enrolled per compound. Household surveys and sample collection were conducted by trained field officers and assistants. Surveys with the primary caregiver of an under-5 child recorded household demographics; water, sanitation, and hygiene access and practices; and animal husbandry practices. A household soil and stored water sample was then collected from each household. Free chlorine levels were measured in a subset of stored water samples (47.5%; n = 57) to characterize chlorine levels in villages (see Supplementary Information for additional sample collection details). Samples were transported to Kenya Medical Research Institute (KEMRI) on ice and were processed within 6 hours of collection. *E. coli* were cultured on Tryptone Bile X-glucuronide (TBX) agar by incubation for 18 hours at 44.5 °C. Specifically, 100 mL of undiluted water samples were membrane filtered and incubated on TBX. Bacteria were eluted from approximately 5.0 g of each soil sample using 50 mL of distilled water and vortexing vigorously for 2 minutes. A 10^-2^ dilution was prepared for each eluted sample; 10 mL of the eluant was membrane filtered and incubated on TBX. Following incubation, we enumerated *E. coli* from TBX plates with up to 500 colony-forming units (CFU). Written informed consent was obtained from all participants, and the study was approved by the KEMRI Scientific and Ethics Review Unit (12/3823) and Tufts Health Sciences Institutional Review Board (13205). The research permit was granted by the Kenyan government through the National Commission for Science, Technology and Innovation (19/2219/29298).

We conducted multivariable linear regressions to determine whether household poultry ownership was associated with (1) *E. coli* prevalence in household stored water and (2) log_10_ *E. coli* concentration in household soil (CFU per dry gram). In a secondary analysis, we used the number of poultry owned by the household as our independent variable. For both analyses, we pre-screened the following covariates, which had <10% missingness and >5% prevalence, for association with each outcome: subcounty; number of households in the compound; number of household residents; asset ownership (television, bike, gas cooker); whether the respondent completed primary education; whether the household had a finished floor (concrete, tiles), obtained water from a piped source, or reported treating stored water; and whether the stool of at least one under-5 child in the household was safely disposed the last time the child passed stool. We retained covariates with p ≤ 0.1 in the models. We modeled *E. coli* prevalence using Poisson regression and log_10_ *E. coli* concentration using a generalized linear model. All analyses were conducted in R (version 3.6.2).

Forty-nine percent (n = 59) of households owned poultry (primarily chickens); ownership of other animals (ruminants, dogs, cats) was less common (Table 1). Poultry-owning households owned 8 poultry on average (median: 4 poultry). Households owning and not owning poultry were similar across multiple sociodemographic indicators. Most households had a concrete floor (88.3%), obtained water from a piped source (74.2%), and had access to an improved latrine (95.0%). About half of households reported safely disposing the stool of young children. Alternatively, poultry-owning households had higher ownership of assets compared to non-poultry-owning households. Additionally, poultry-owning households more commonly had feces present around the household soil sampling area—although feces were still observed around some non-poultry-owning households.

**Table 1.** Household characteristics by household poultry ownership.

|  | Percent of Households (n)<sup>a</sup> |  |  |
| --- | --- | --- | --- |
|  | Poultry<br>(N = 59) | No Poultry<br>(N = 61) | Overall<br>(N = 120) |
| Respondent's highest level of completed education |  |  |  |
| Primary | 83.1 (49) | 78.7 (48) | 80.8 (97) |
| Secondary | 28.8 (17) | 32.8 (20) | 30.8 (37) |
| Households in compound |  |  |  |
| Median (mean) | 9 (14.0) | 10 (11.8) | 10 (12.9) |
| Residents in household |  |  |  |
| Median (mean) | 5 (5.4) | 4 (4.6) | 5 (5.0) |
| Main material of floor |  |  |  |
| Earth/dung | 8.5 (5) | 6.6 (4) | 7.5 (9) |
| Concrete | 89.8 (53) | 86.9 (53) | 88.3 (106) |
| Tiles | 1.7 (1) | 4.9 (3) | 3.3 (4) |
| No data | 0.0 (0) | 1.6 (1) | 0.8 (1) |
| Household Assets |  |  |  |
| Bicycle | 15.3 (9) | 11.5 (7) | 13.3 (16) |
| Car | 1.7 (1) | 0.0 (0) | 0.8 (1) |
| Clock | 25.4 (15) | 14.8 (9) | 20.0 (24) |
| Electricity | 100.0 (59) | 95.1 (58) | 97.5 (117) |
| Gas cooker | 78.0 (46) | 63.9 (39) | 70.8 (85) |
| Mobile phone | 98.3 (58) | 93.4 (57) | 95.8 (115) |
| Motorcycle | 8.5 (5) | 8.2 (5) | 8.3 (10) |
| Radio | 83.1 (49) | 68.9 (42) | 75.8 (91) |
| Stove | 69.5 (41) | 80.3 (49) | 75.0 (90) |
| Television | 89.8 (53) | 80.3 (49) | 85.0 (102) |
| Source water source |  |  |  |
| Borehole | 20.3 (12) | 21.3 (13) | 20.8 (25) |
| Piped water into yard/plot | 10.2 (6) | 3.3 (2) | 6.7 (8) |
| Piped water outside the compound | 64.4 (38) | 70.5 (43) | 67.5 (81) |
| Tanker truck | 5.1 (3) | 1.6 (1) | 3.3 (4) |
| No data | 0.0 (0) | 3.3 (2) | 1.7 (2) |
| Stored water treated<sup>b</sup> |  |  |  |
| Yes | 15.3 (9) | 18.0 (11) | 16.7 (20) |
| No | 83.1 (49) | 78.7 (48) | 80.8 (97) |
| No data or don't know/refused | 1.7 (1) | 3.3 (2) | 2.5 (3) |
| Detectable free chlorine in stored water<sup>c</sup> |  |  |  |
| Yes | 8.5 (5) | 6.6 (4) | 7.5 (9) |
| No | 40.7 (24) | 39.3 (24) | 40.0 (48) |
| No data | 50.8 (30) | 54.1 (33) | 52.5 (63) |
| Toilet owner |  |  |  |
| Household only | 10.2 (6) | 1.6 (1) | 5.8 (7) |
| Household shared | 50.8 (30) | 59.0 (36) | 55.0 (66) |
| Someone inside the compound | 6.8 (4) | 3.3 (2) | 5.0 (6) |
| Someone outside the compound | 10.2 (6) | 14.8 (9) | 12.5 (15) |
| Public | 20.3 (12) | 18.0 (11) | 19.2 (23) |
| No data | 1.7 (1) | 3.3 (2) | 2.5 (3) |
| Improved latrine<sup>d</sup> |  |  |  |
| Yes | 94.9 (56) | 95.1 (58) | 95.0 (114) |
| No | 3.4 (2) | 0.0 (0) | 1.7 (2) |
| No data | 1.7 (1) | 4.9 (3) | 3.3 (4) |
|  | Poultry<br>(N = 59) | No Poultry<br>(N = 61) | Overall<br>(N = 120) |
| Safely disposed stool last (under 5 years)<sup>e</sup> |  |  |  |
| Yes | 52.5 (31) | 52.5 (32) | 52.5 (63) |
| No | 47.5 (28) | 47.5 (29) | 47.5 (57) |
| Household ownership of other animals |  |  |  |
| Ruminants<sup>f</sup> | 3.4 (2) | 0.0 (0) | 1.7 (2) |
| Dogs | 10.2 (6) | 0.0 (0) | 5.0 (6) |
| Cats | 27.1 (16) | 4.9 (3) | 15.8 (19) |
| Feces around the household soil sampling area<sup>g</sup> |  |  |  |
| Yes | 32.2 (19) | 8.2 (5) | 20.0 (24) |
| No | 67.8 (40) | 91.8 (56) | 80.0 (96) |

Note: Poultry ownership (chickens, ducks, turkey, and other poultry) was similar between both subcounties (31/64 = 48.4% of households in Kibera owned poultry and 28/56 = 50.0% of households in Dagoretti South owned poultry).
<sup>a</sup> For number of households in compound and number of residents in household, median (mean) are reported instead.
<sup>b</sup> The treatment methods included in the survey were chlorine dispenser, waterguard/bottled chlorine, boiling, sieving, other types of filters (e.g., ceramic), solar disinfection, letting water stand and settle, biosand filter, Vestergaard Frandsen lifestraw, coagulant, Pur, Aquatabs, and chlorine. Among these treatment methods, only waterguard/bottled chlorine, boiling, other filter (e.g., ceramic), and/or Aquatabs were reported by households.
<sup>c</sup> Detectable free chlorine was defined as $\geq 0.1$ mg/L. Chlorine was only tested from water samples in a small subset of households (n = 57 households for stored water). Refer to the Supplementary Information for more details.
<sup>d</sup> Improved latrines were either composting latrines or latrines with a slab and/or vent. All latrines with a vent had a slab and all composting latrines had a slab.
<sup>e</sup> Stool was safely disposed the last time the child passed stool if the child used a latrine or their stool was rinsed into a latrine. For households with more than one child under 5 years, this variable was 'yes' if at least one child's stool was safely disposed.
<sup>f</sup> Ruminants include cows, goats, and sheep.
<sup>g</sup> Feces types observed include poultry, dog/cat, and human (poultry feces were the most common type of feces present).

The following statistics pertain only to households in poultry-owning compounds (55.0%; n = 66) (Table 2). The most common purposes of poultry rearing were meat, eggs, and income generation. Poultry were most commonly free-ranging and/or fed food scraps and maize; poultry were less commonly fed commercial feed. About half of households reported giving poultry medicine in the past, with antibiotics and vaccines being the most common types of medications administered. Poultry entered more households in Kibera (63.9%; n = 23) than Dagoretti South (53.3%; n = 16), which is less densely populated than Kibera. Among these households, more households in Kibera (69.6%; n = 16) kept poultry in the sleeping quarters of under-5 children compared to Dagoretti South (31.3%; n = 5). Rearing practices of allowing chickens to roam freely around both the household and compound were similar in peri-urban Kisumu, Kenya^9^, suggesting that our study site was representative of other urban sites in Kenya. The household respondent cared for poultry in 84.9% of compounds; other adults and older children from within and outside the household also participated in poultry caretaking. Other studies also observed that women and children often care for poultry^10,11^, suggesting that these two populations may experience a higher risk of exposure to poultry feces.

**Table 2.**
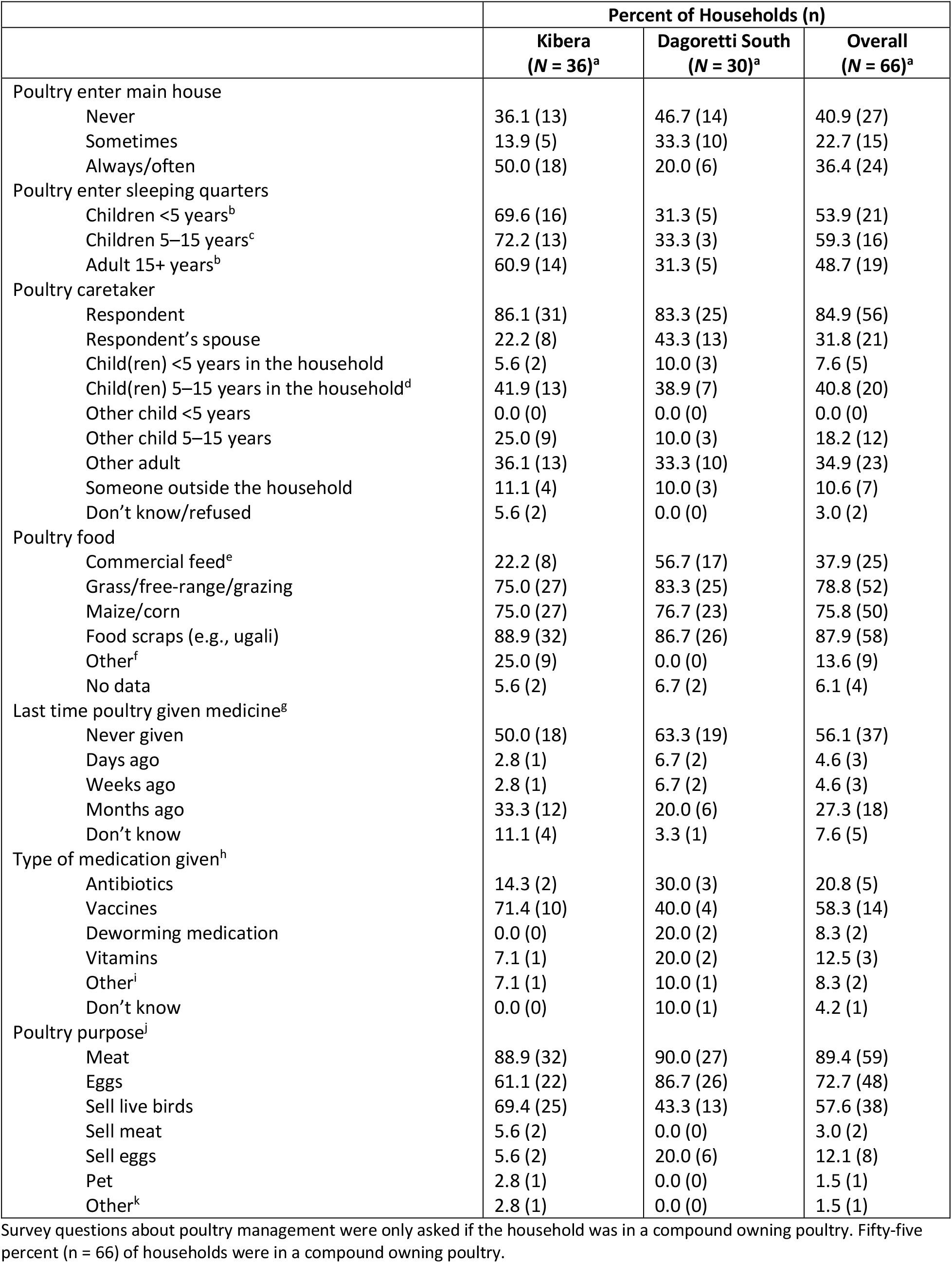
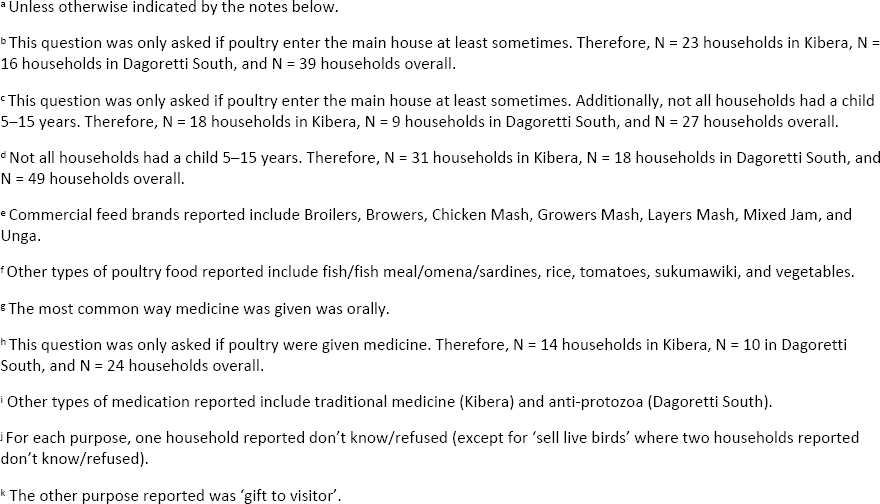
Household poultry management by subcounty.

|  | Percent of Households (n) |  |  |
| --- | --- | --- | --- |
|  | Kibera<br>(N = 36)<sup>a</sup> | Dagoretti South<br>(N = 30)<sup>a</sup> | Overall<br>(N = 66)<sup>a</sup> |
| Poultry enter main house |  |  |  |
| Never | 36.1 (13) | 46.7 (14) | 40.9 (27) |
| Sometimes | 13.9 (5) | 33.3 (10) | 22.7 (15) |
| Always/often | 50.0 (18) | 20.0 (6) | 36.4 (24) |
| Poultry enter sleeping quarters |  |  |  |
| Children <5 years<sup>b</sup> | 69.6 (16) | 31.3 (5) | 53.9 (21) |
| Children 5–15 years<sup>c</sup> | 72.2 (13) | 33.3 (3) | 59.3 (16) |
| Adult 15+ years<sup>b</sup> | 60.9 (14) | 31.3 (5) | 48.7 (19) |
| Poultry caretaker |  |  |  |
| Respondent | 86.1 (31) | 83.3 (25) | 84.9 (56) |
| Respondent's spouse | 22.2 (8) | 43.3 (13) | 31.8 (21) |
| Child(ren) <5 years in the household | 5.6 (2) | 10.0 (3) | 7.6 (5) |
| Child(ren) 5–15 years in the household<sup>d</sup> | 41.9 (13) | 38.9 (7) | 40.8 (20) |
| Other child <5 years | 0.0 (0) | 0.0 (0) | 0.0 (0) |
| Other child 5–15 years | 25.0 (9) | 10.0 (3) | 18.2 (12) |
| Other adult | 36.1 (13) | 33.3 (10) | 34.9 (23) |
| Someone outside the household | 11.1 (4) | 10.0 (3) | 10.6 (7) |
| Don't know/refused | 5.6 (2) | 0.0 (0) | 3.0 (2) |
| Poultry food |  |  |  |
| Commercial feed<sup>e</sup> | 22.2 (8) | 56.7 (17) | 37.9 (25) |
| Grass/free-range/grazing | 75.0 (27) | 83.3 (25) | 78.8 (52) |
| Maize/corn | 75.0 (27) | 76.7 (23) | 75.8 (50) |
| Food scraps (e.g., ugali) | 88.9 (32) | 86.7 (26) | 87.9 (58) |
| Other<sup>f</sup> | 25.0 (9) | 0.0 (0) | 13.6 (9) |
| No data | 5.6 (2) | 6.7 (2) | 6.1 (4) |
| Last time poultry given medicine<sup>g</sup> |  |  |  |
| Never given | 50.0 (18) | 63.3 (19) | 56.1 (37) |
| Days ago | 2.8 (1) | 6.7 (2) | 4.6 (3) |
| Weeks ago | 2.8 (1) | 6.7 (2) | 4.6 (3) |
| Months ago | 33.3 (12) | 20.0 (6) | 27.3 (18) |
| Don't know | 11.1 (4) | 3.3 (1) | 7.6 (5) |
| Type of medication given<sup>h</sup> |  |  |  |
| Antibiotics | 14.3 (2) | 30.0 (3) | 20.8 (5) |
| Vaccines | 71.4 (10) | 40.0 (4) | 58.3 (14) |
| Deworming medication | 0.0 (0) | 20.0 (2) | 8.3 (2) |
| Vitamins | 7.1 (1) | 20.0 (2) | 12.5 (3) |
| Other<sup>i</sup> | 7.1 (1) | 10.0 (1) | 8.3 (2) |
| Don't know | 0.0 (0) | 10.0 (1) | 4.2 (1) |
| Poultry purpose<sup>j</sup> |  |  |  |
| Meat | 88.9 (32) | 90.0 (27) | 89.4 (59) |
| Eggs | 61.1 (22) | 86.7 (26) | 72.7 (48) |
| Sell live birds | 69.4 (25) | 43.3 (13) | 57.6 (38) |
| Sell meat | 5.6 (2) | 0.0 (0) | 3.0 (2) |
| Sell eggs | 5.6 (2) | 20.0 (6) | 12.1 (8) |
| Pet | 2.8 (1) | 0.0 (0) | 1.5 (1) |
| Other<sup>k</sup> | 2.8 (1) | 0.0 (0) | 1.5 (1) |

Survey questions about poultry management were only asked if the household was in a compound owning poultry. Fifty-five percent (n = 66) of households were in a compound owning poultry.
<sup>a</sup> Unless otherwise indicated by the notes below.
<sup>b</sup> This question was only asked if poultry enter the main house at least sometimes. Therefore, N = 23 households in Kibera, N = 16 households in Dagoretti South, and N = 39 households overall.
<sup>c</sup> This question was only asked if poultry enter the main house at least sometimes. Additionally, not all households had a child 5–15 years. Therefore, N = 18 households in Kibera, N = 9 households in Dagoretti South, and N = 27 households overall.
<sup>d</sup> Not all households had a child 5–15 years. Therefore, N = 31 households in Kibera, N = 18 households in Dagoretti South, and N = 49 households overall.
<sup>e</sup> Commercial feed brands reported include Broilers, Browers, Chicken Mash, Growers Mash, Layers Mash, Mixed Jam, and Unga.
<sup>f</sup> Other types of poultry food reported include fish/fish meal/omena/sardines, rice, tomatoes, sukumawiki, and vegetables.
<sup>g</sup> The most common way medicine was given was orally.
<sup>h</sup> This question was only asked if poultry were given medicine. Therefore, N = 14 households in Kibera, N = 10 in Dagoretti South, and N = 24 households overall.

*E. coli* was detected in 36.7% (n = 44) of stored water samples (range: 100–50,119 CFU/100 ml) (Figure 1). The prevalence of *E. coli* in water was not higher in poultry-owning households than non-poultry-owning households (40.7% vs 32.8%; χ^2^ = 0.3; p = 0.6). *E. coli* was detected in 85.0% (n = 102) of household soil samples (range: 50– 501,187 CFU/dry g soil), and the prevalence of *E. coli* in soil was significantly higher in poultry-owning than non-poultry-owning households (94.9% vs 75.4%; Fisher’s exact; p < 0.01).

**Figure 1.**
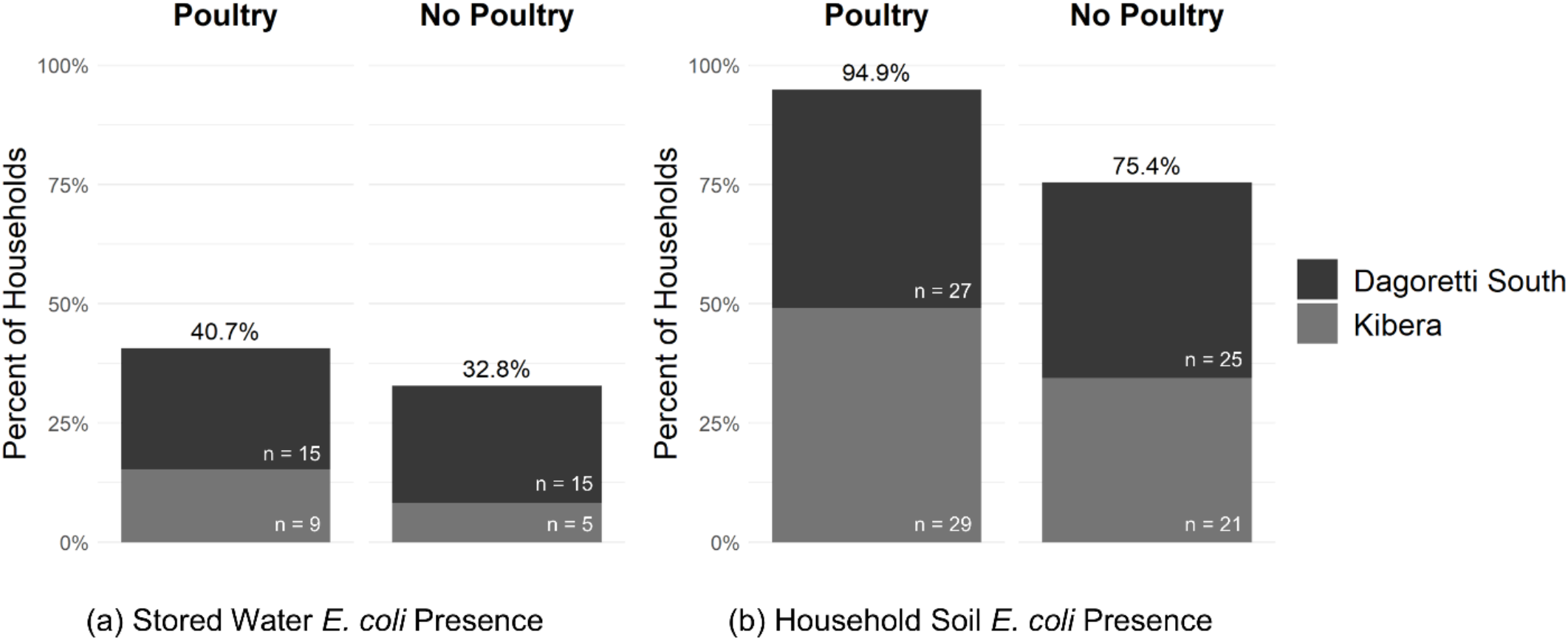
*E. coli* presence in (a) household stored water and (b) household soil samples stratified by household poultry ownership. Forty-nine percent of households (n = 59) owned poultry (n = 31 in Kibera, n = 28 in Dagoretti South). Overall, *E. coli* was detected in 36.7% (n = 44) of household stored water samples; 3.3% of households (n = 4) did not have stored water *E. coli* data. In household soil samples, *E. coli* was detected in 85.0% (n = 102) of households overall.

Adjusting for significant covariates, household poultry ownership was not associated with *E. coli* prevalence in household stored water but was associated with increased log_10_ *E. coli* concentration in household soil (Table S1). Poultry-owning households had a 0.67 (95% confidence interval: 0.27–1.07) log_10_ CFU/dry g increase in soil *E. coli* compared to non-poultry-owning households. The number of poultry owned by a household was also positively associated with *E. coli* concentration in household soil; there was a 0.03 (95% confidence interval: 0.00–0.05) log_10_ CFU/dry g increase in soil *E. coli* for each additional poultry animal owned. Similarly, a previous study in Kibera found that ownership or presence of chickens in the compound was associated with increased *Campylobacter jejuni*, an enteric pathogen, in household soil.^12^ Researchers also observed a correlation between animal ownership and soil *E. coli* in dense communities of Harare, Zimbabwe.^13^

Our study has limitations. Chlorine was detected in some household stored water, but we could not include free chlorine residual as a covariate in our models because chlorine measurements were only collected from a small subset of households. Additionally, we were limited by sample size. Future studies with a greater sample size should investigate the relationship between *E. coli* contamination and indicators other than poultry ownership (e.g., day and nighttime corralling practices, animal feces disposal practices). Finally, we only measured FIB instead of actual pathogens. In urban settings where animals live near humans, investigating animal management practices may be especially important for understanding household- and community-level transmission of zoonotic enteropathogens.

Poultry ownership in dense settings may promote zoonotic disease transmission. In urban Kenya, household soil may be an important transmission pathway for poultry-associated fecal contamination. Finished flooring (concrete, tiles) is one intervention that has been hypothesized to disrupt transmission of fecal contamination from soil. However, the prevalence of finished flooring was high in our study, suggesting that areas immediately outside of the living quarters where people spend a lot of time (e.g., cleaning, playing) could still be an important place for human exposure to poultry fecal contamination. Additional research is needed to understand how animal husbandry practices influence environmental contamination in urban settlements in order to inform potential interventions for disrupting zoonotic disease transmission in low-income households.

## Data Availability

All data produced in the present work are contained in the manuscript.

## Acknowledgments

We sincerely thank the study participants for their gracious contributions to this research. We thank Anne Okello, Fredrick Okuku Owuor, Eric Kipchirchir Tanui, Glory Wairimu, and Henry Emonje Mukabwa for sample and data collection, Frank Odhiambo for coordinating study logistics, Maya Nadimpalli for assistance with developing protocols, survey instruments, and coordinating study logistics, Tim Julian for assistance with developing *E. coli* isolation methods, Gretchen Walch, Deeksha Bathini, Gracie Hornsby, and Jeremy Lowe for their contributions to sample processing and survey data review, and the many KEMRI attachments and interns for their willingness to learn and assist with sample processing.

## Authors’ Contact Information

Elana Chan: Department of Civil and Environmental Engineering, Stanford University, Stanford, CA, USA

Jenna Swarthout: Department of Civil and Environmental Engineering, Tufts University, Medford, MA, USA

Benard Chieng: Kenya Medical Research Institute (KEMRI), Mbagathi Road, P.O. Box 54840-00200 Nairobi, Kenya

Sylvie Araka: Kenya Medical Research Institute (KEMRI), Mbagathi Road, P.O. Box 54840-00200 Nairobi, Kenya

John Mboya: Department of Civil and Environmental Engineering, University of California, Berkeley, Berkeley, CA 94720

Christine Imali: Department of International Development, London School of Economics and Political Science, London, UK

Angela Harris: Department of Civil, Construction, and Environmental Engineering. North Carolina State University, Raleigh, NC, 27695

Sammy Njenga: Kenya Medical Research Institute (KEMRI), Mbagathi Road, P.O. Box 54840-00200 Nairobi, Kenya

Amy Pickering: Department of Civil and Environmental Engineering, University of California, Berkeley, Berkeley, CA 94720

