## Supplemental Information for "Poultry Ownership in Urban Kenya is Associated with Increased Fecal Contamination in Household Soil"

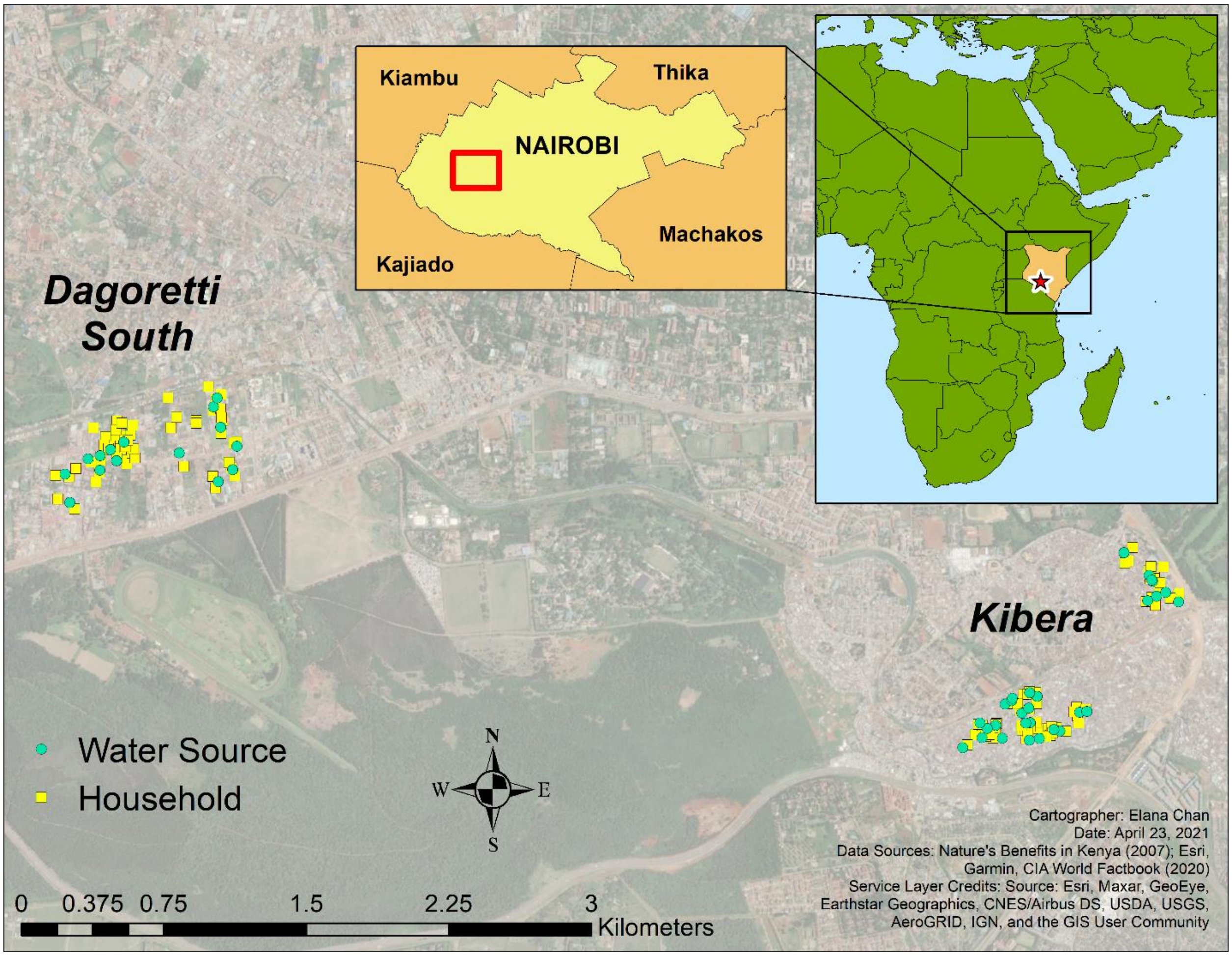


Figure S1. Location of households and water sources in the two subcounties in Nairobi County, Kenya (Dagoretti South and Kibera).

**Additional Sample and Survey Data Collection Details**

In addition to a household stored water and soil sample, a source water and water source soil sample were also collected for each household. If more than one household used the same water source, only one water and soil sample was collected per water source. Water and soil data from the water source were then linked to all households using that water source.

Free chlorine levels were measured in stored and source water samples from 47.5% (n = 57) and 27.5% (n = 33) of households, respectively. Water samples collected prior to July 26, 2019, were not treated with sodium thiosulfate which is why free chlorine levels were not measured in water samples from all households. This study was designed for detecting helminths, but after a couple weeks of detecting low *E. coli* prevalence in the water samples, we realized in early July that chlorine levels must be higher in the study area than expected. Because we were leveraging the samples from this study for *E. coli* culturing, we ordered whirlpak bags with sodium thiosulfate. We started free chlorine measurements on July 11, but due to logistical concerns, we decided not to revisit household from which we had already collected water samples. Among households with stored and source water chlorine measurements, 7.5% (n = 9) and 8.3% (n = 10) of households, respectively, had detectable free chlorine (≥ 0.1 mg/L).

For our main statistical analyses, we ultimately decided not to analyze *E. coli* in source water and water source soil samples as outcome variables because water source points were often located outside of a household's compound, and poultry ownership at the household level was our main exposure variable of interest. For reference though, 5.0% (n = 6) of households had *E. coli* detected in source water (range: 100–708 CFU/100 ml); 15.0% (n = 18) of household did not have source water *E. coli* data. Seventy-five percent (n = 90) of households had *E. coli* detected in water source soil (range: 245–537,032 CFU/dry g soil); 10.0% (n = 12) of household did not have water source soil *E. coli* data.

Survey data about animal husbandry practices were also collected for dogs, but dog ownership was less common; only 20.8% (n = 25) of households were in a dog-owning compound. Based on this small subpopulation, dogs entered 8.0% (n = 2) of households. Ninety-two percent (n = 23) of households reared dogs for security; only one household kept dogs as pets; and no households reared dogs for hunting, herding, or to sell.

Table S1. Regression output

| | Main Exposure | | | |
|---|---|---|---|---|
| | Poultry Ownership (yes/no) | | Poultry Ownership (# owned)[a] | |
| (1) Outcome: *E. coli* Presence in Household Stored Water (N = 115) | | | | |
| Variable | PR (95% CI) | p-value | PR (95% CI) | p-value |
| **Poultry Ownership** | 1.10 (0.60, 2.03) | 0.75 | 1.01 (0.98, 1.04) | 0.45 |
| Subcounty (Kibera) | 0.47 (0.24, 0.91) | 0.03 | 0.48 (0.24, 0.92) | 0.03 |
| Households | 1.01 (0.99, 1.03) | 0.34 | 1.01 (0.99, 1.03) | 0.32 |
| Gas Cooker | 1.41 (0.67, 3.37) | 0.40 | 1.38 (0.65, 3.30) | 0.43 |
| (2) Outcome: Log10 *E. coli* Concentration in Household Soil (N = 104) | | | | |
| Variable | Coeff (95% CI) | p-value | Coeff (95% CI) | p-value |
| **Poultry Ownership** | 0.67 (0.27, 1.07) | <0.01 | 0.03 (0.00, 0.05) | 0.05 |
| Piped | 0.38 (-0.14, 0.90) | 0.15 | 0.41 (-0.13, 0.94) | 0.14 |

*E. coli* presence in household stored water was modeled using a Poisson regression model. Log10 *E. coli* concentration in household soil was modeled using a generalized linear regression model. *E. coli* concentrations were originally calculated in colony forming units per dry gram of household soil. Pre-screened covariates with $p \leq 0.1$ were retained in the models.

[a] If the number of poultry animals for a given poultry type (e.g., chickens) was unknown, the household was assumed to own zero poultry for the poultry type when calculating total number of poultry owned by the household (i.e., counts may be underestimated).

CI = confidence interval

Coeff = regression coefficient

N = number of complete observations

PR = prevalence ratio

Poultry Ownership: When modeling poultry ownership as a binary (yes/no) exposure variable, the variable was dummy coded (1 = household owns at least one poultry animal, 0 = household owns zero poultry animals). When modeling poultry ownership as a discrete exposure variable (# owned), the variable value was the number of poultry owned by the household.

Subcounty: 1 = Kibera, 0 = Dagoretti South

Households: Number of households in the household's compound. One household reported 100 households in its compound; this observation was deleted when running the regression model.

Gas cooker: 1 = household has a gas cooker, 0 = household does not have a gas cooker

Piped: 1 = household obtains water from a piped source, 0 = household does not obtain water from a piped source
